# Assessment of N95 and K95 respirator decontamination: fiber integrity, filtration efficiency, and dipole charge density

**DOI:** 10.1101/2020.07.07.20148551

**Authors:** Wonjun Yim, Diyi Cheng, Shiv Patel, Rui Kui, Ying Shirley Meng, Jesse V. Jokerst

## Abstract

Personal protective equipment (PPE) including N95 respirators are critical for persons exposed to SARS-CoV-2. KN95 respirators and N95 decontamination protocols have been described as solutions to a lack of such PPE. However, there are a few materials science studies that characterize the charge distribution and physical changes accompanying disinfection treatments particularly heating. Here, we report the filtration efficiency, dipole charge density, and fiber integrity of pristine N95 and KN95 respirators before and after various decontamination methods. We found that the filter layer of N95 is 8-fold thicker than that of KN95, which explains its 10% higher filtration efficiency (97.03 %) versus KN95 (87.76 %) under pristines condition. After 60 minutes of 70 °C treatment, the filtration efficiency and dipole charge density of N95 became 97.16% and 12.48 μC/m^2^, while those of KN95 were 83.64% and 1.48 μC/m^2^; moreover, fit factor of N95 was 55 and that of KN95 was 2.7. In conclusion, the KN95 respirator is an inferior alternative of N95 respirator. In both systems, a loss of electrostatic charge does not directly correlate to a decrease in performance.

---

The ongoing Coronavirus Disease 2019 (COVID-19) pandemic has had a major impact on human health and society with a mortality rate apparently higher than influenza.^1^ COVID-19 results from infection by the severe acute respiratory syndrome coronavirus 2 (SARS-CoV-2) in which the spike (S) protein on the SARS-CoV-2 plays a key role in mediating viral entry into the human cell.^2^ The main route of transmission between humans is likely aerosols and droplets.^3^ When an infected person coughs, sneezes, or speaks, the virus will be excreted and dissolved with the aerosol forming the viral-containing droplets (>5 to 10 *μ*m) or viral aerosols (≤5 *μ*m) that can remain and travel in the air.^4^

N95 respirators have been used to protect wearers against such viral aerosols and droplets. They have at least 95% filtration efficiency for NaCl particles sized 0.1 to 0.3 *μ*m with even higher filtration efficiency at higher particle size (approximately 99.5% or higher for 0.75 *μ*m particles).^5^ Hence, N95 respirators offer excellent protection when they seal tightly over the face. Filter fabric are made of, nylon, cotton, polyester (PE) and polypropylene (PP).^6-7^ Nylon filter has a good resistance to rubbing and cotton filter are friendly to environment. PE filter has good acid-resistance and excellent durability against elevated temperature up to 150 °C.^8^ PP filter is the lightest weight among synthetic fabrics and has good resistance against acids and alkalis.^8^ Nonwoven PP fabric is composed of a random fibrous web in which individual fibers are bound together in a random arrangement—thus, particles inhaled with air interact with the fiber and adhere efficiently.^9^

Spin-bonding and melt-blowing are two key manufacturing processes for fabricating non-woven PP fabric.^10^ The diameter of the fiber obtained from spun-bond process is thicker than melt-blown process.^11^ Hence, spun-bond fiber has been used as the outer and inner layer of respirators to provide mechanical support for other structural or filter layers leading to enhanced durability.^12^ According to the filtration theory of elementary fibers,^13^ the degree of packing density of fiber and electrostatic charge determines the filtration efficiency of nonwoven fabric. Therefore, a high packing density of melt-blown PP fiber (2 m^2^/g)—at least 10-fold larger than spun-bond (0.2 m^2^/g)—plays a key role in the filtration performance.^14^ In addition to high packing density, filtration efficiency can be dramatically improved by electrostatic charges imparted on the filter fibers.^15^ An electrostatic field applied during manufacturing process is a common way to induce electrostatic charges within non-woven PP fibers used as a filter layer of the respirators.^16^ This field leaves PP microfibers with dipole charges on their surface thus improving their filtration efficiency.^17-18^

Shortages of PPE such as medical masks and N95 respirators for healthcare workers have been widely reported and have become top concerns for hospitals.^19^ Although there has been nanoparticle developments for treating and imaging of viral infection,^20-21^ demand-pull inflation of N95 respirators leads to supply limits and counterfeit N95 respirators are recently causing additional risks to healthcare workers.^22^ Thus, a variety of decontamination methods have been studied to reuse N95 respirators. Vaporized hydrogen peroxide (VHP), 70 °C dry-heat, ultraviolet light (UV), and 70% ethanol have all been described and can inactivate SARS-CoV-2.^23^ Although the VHP method is a well-known sterilization technology^24^ that has been approved by FDA,^25^ it needs complicated equipment and a trained technician. The ethanol method is found to damage N95 respirators after first cycle of decontamination, and UV-radiation has limited penetration through the multiple layers of the respirator.^23^ Hence, dry-heat has emerged as a simple and promising decontamination method;^26-27^ it uniformly disinfects respirators with good scalability. However, an understanding of the physical changes induced by these treatments remains incomplete.

We evaluate the effect of decontamination on respirators and include a special emphasis on KN95 respirators as a potential alternative to N95 respirators from a performance and material science properties. Recent works^7, 23, 26-29^ focused nearly exclusively on the performance of the respirators, i.e., their filtration performance. Here, for the first time, we carefully studied not only filtration efficiency but also dipole charge density which is related with electrostatic filtration but has yet been investigated during decontamination and fiber integrity of pristine N95 and KN95 respirators as well as these same respirators after various decontamination methods.

## Results and Discussion

### Structural components of N95 and KN95 respirator

KN95 respirator which follows Chinese standards^30^ is described as having similar filtration performance as N95 but are not NIOSH approved. We disassembled N95 and KN95 to compare their structural elements. The N95 respirator had three major layers: outer layer, filter layer, and inner layer (**Figure 1A**) while the KN95 respirator had four major layers: outer layer, filter layer, middle layer and inner layer (**Figure 1B**). Both respirators used spun-bond PP fabric for the outer/inner layer and melt-blown PP fabric for the filter layer. The middle layer of KN95 was composed of cotton fibers in which two components formed core-shell fiber structure (**Figure 1B**).

**Figure 1.**
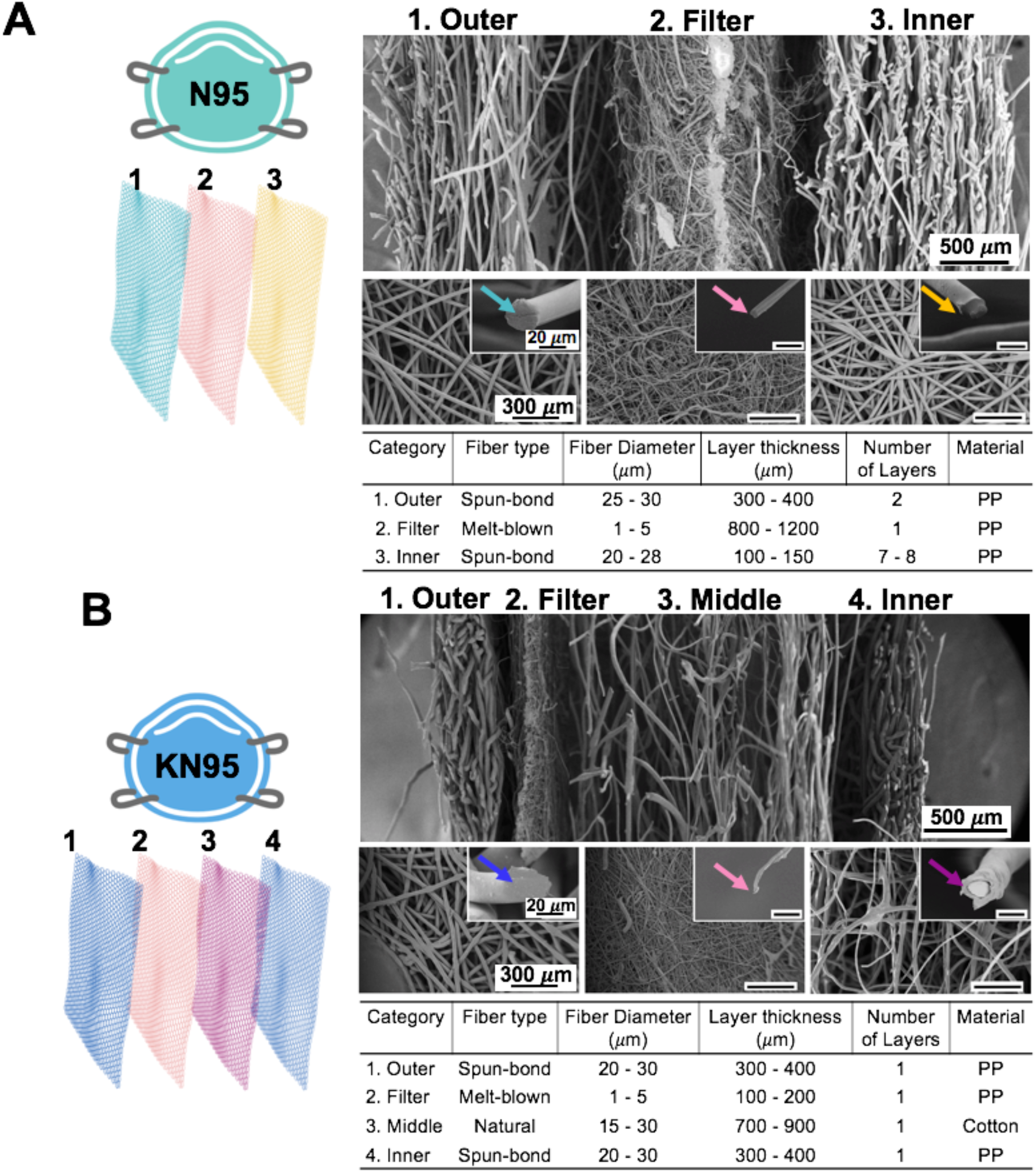
Structural components of N95 and KN95 respirators. SEM image shows lateral and front view of multiple layers in (A) N95 and (B) KN95 respirators. (A) N95 is composed of outer layer (spun-bond PP), filter layer (melt-blown PP), and inner layer (spun-bond PP). Green, pink, and yellow arrows point to individual fiber in the outer, filter, and inner layer. (B) KN95 respirator is composed of outer layer (spun-bond PP), filter layer (melt-blown PP), middle layer (cotton), and inner layer that is identical to the outer layer. Blue, pink, and purple arrows point to individual fiber in the outer, filter, and middle layer. Table includes detailed information for each layer: fiber type, fiber diameter, layer thickness, number of layers, and material.

The fiber diameter of the outer layer in the N95 was 27.07 μ*m* ± 3.64 μ*m* and that of KN95 was 26.07 μ*m* ± 3.63 μ*m*. The fiber diameter of filter layer in the N95 was 2.79 μ*m* ± 0.95 μ*m* and that of KN95 was 3.23 μ*m* ± 1.28 μ*m*. Hence, the filter layer on both respirators have good mechanical filtration since smaller fibers make smaller pore-size area.^31^ Interestingly, the layer thickness of the filter layer in the N95 was 8-fold thicker than KN95 (**Figure 1**). Instead, the KN95 had a supplementary middle layer. While the KN95 used a single thick layer of spun-bond PP fabric for the inner layer, the N95 used multiple thin layers of spun-bond PP fabric. Those structural differences especially for the thickness of filter layer explained the higher filtration efficiency of the N95 than the KN95’s (**Figure 2D and Figure 2I**).

**Figure 2.**
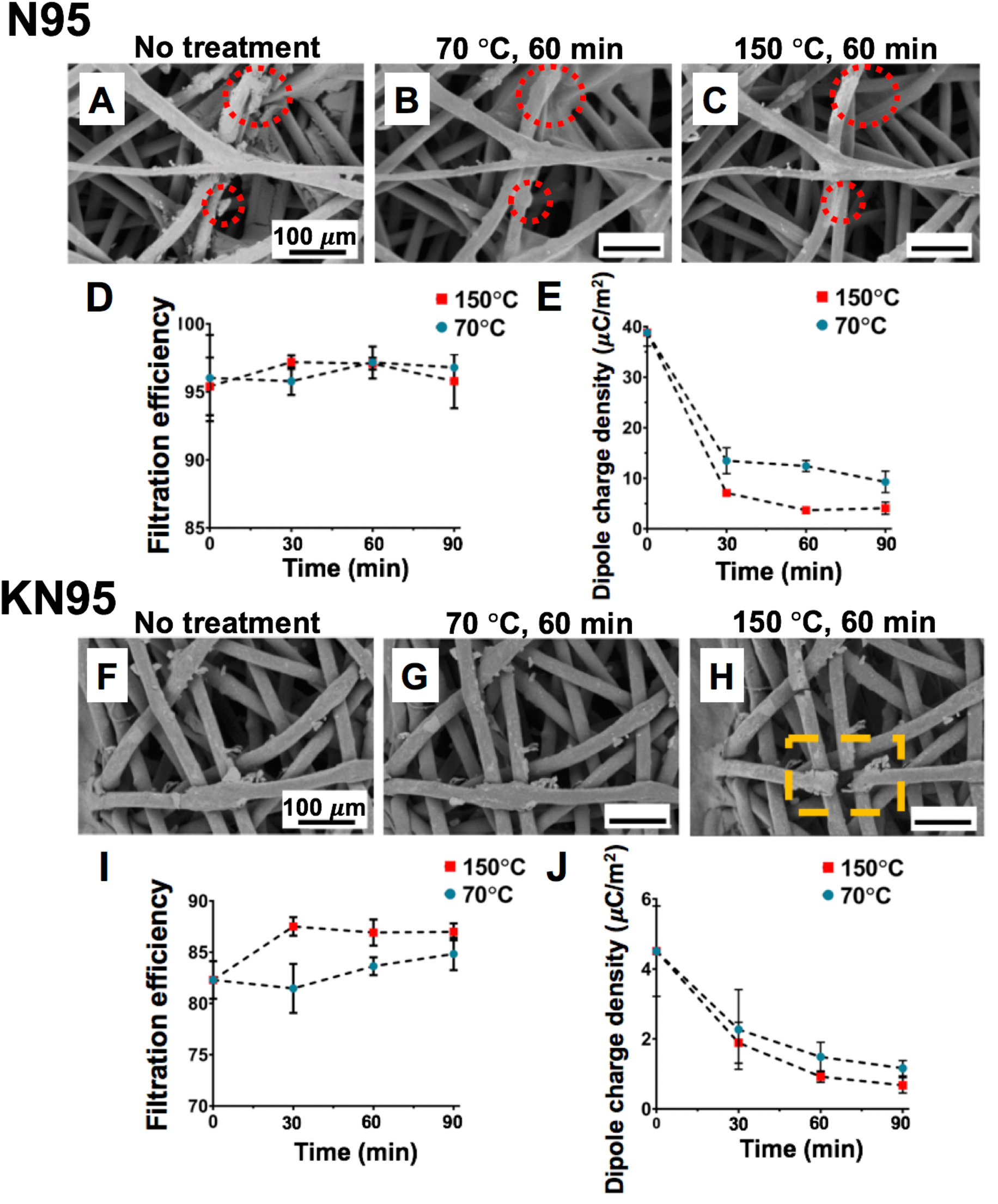
Decontamination of N95 and KN95 respirator. (A−C) Dust removal is seen at the outer layer of N95 respirator. Red circle shows dust particles disappear after heat treatment. (D) filtration efficiencies of N95 after 70 °C and 150 °C treatment (E) Dipole charge densities of N95 decrease with heat treatment. (F−G) Few particles are removed at the outer layer of KN95 after 70 °C treatment. (H) Fiber fracture occurs at 150 °C treatment. Orange rectangle shows the fracture happens at the outer layer of KN95. (I) filtration efficiencies of KN95 after 70 °C and 150 °C treatment. (J) Dipole charge density of KN95 linearly decreases upon heat treatment. Error bars represent the standard deviations of 5 measurements. All scale bars represent 100 μ*m*.

### Decontamination of N95 and KN95 respirator

Since 70°C treatment is validated by the National Institute of Health (NIH) to inactivate the SARS-CoV-2,^23, 32^ contaminated N95 and KN95 respirators were heat-treated in the oven for three cycles (30 minutes/cycle) at 70 °C and 150 °C for a positive control respectively. We cut the respirators into 2 cm squares and put them in the heating oven. After each cycle of 70 °C and 150 °C treatment, SEM images of the outer layer were taken at the same spot to examine the effect of heat treatment on the particle removal for N95 (**Figure 2A – C**) and KN95 (**Figure 2F – H**).

Due to electrostatic interactions between dusts and fabrics,^33^ plenty of small particles were trapped on the spun-bond PP fibers of the outer layer (**Figure 2A and Figure 2D**). Those particles attached on the fibers were removed after the completion of each cycle in which charge dissipation of fibers occurred at high temperature losing electrical entrapment. Some dust on the fiber melted and removed at the 150 °C treatment (**Figure 2C**). However, there were still a number of particles attached on the outer layer in both N95 and KN95 respirator even after 150 °C treatment (**Figure 2C and Figure 2H**). Since filtration efficiencies are a function of the filter layer, the filtration efficiencies of both respirators were improved versus the baseline condition regardless of the number of particles observed in the outer layers. We also found structural instability such as a fracture only in the outer layer of KN95 (**Figure 2H**); there was no such cracking in N95.

We measured the filtration efficiency and dipole charge density of N95 and KN95 respirators after 70 °C and 150 °C treatment to evaluate the effect of dipole charge density toward the filtration efficiency. Although there was a significant drop of dipole charge density of N95 (**Figure 2E**) and KN95 (**Figure 2J**), there was little change in filtration after the heat treatments (**Figure 2D and Figure 2I**). This was because filtration efficiency of respirator was also affected by mechanical filtration that is based on inertia impaction, interception, and diffusion—these are not markedly influenced by charge.^34^ In addition, the increase in filtration efficiency due to electrostatic attraction is most significant for 2 to 100 nm particles^35^ illustrating that filtration efficiency depends on the particle size and air flow in the air.^36^ Thus, we took a SEM image of the N95 sample after filtration test to measure particle size attached on the fibers. The average particle size attached on the fibers was 15.28 μ*m* ± 5.10 μ*m* (**Figure S1**).

### Comparison of N95 and KN95 respirator

We chose to focus on 70 °C treatment which can inactivate SARS-CoV-2 with no damage on the fiber integrity. Filtration efficiencies of N95 respirator after each cycle of 70 °C treatment remained over 95 % while that of KN95 was around 83 % (**Figure 3A**). At least 10 % gap of the filtration efficiency between N95 and KN95 might result from the larger thickness of filter layer in N95 respirator (**Figure 1**). In addition, the fit factor of KN95 stayed close to 2 even after the heat treatment, illustrating KN95 is loose-fitting to the face, while fit factor of N95 was 10-fold higher than KN95’s which means it was tight-fitting to the face. (**Figure 3B**)

**Figure 3.**
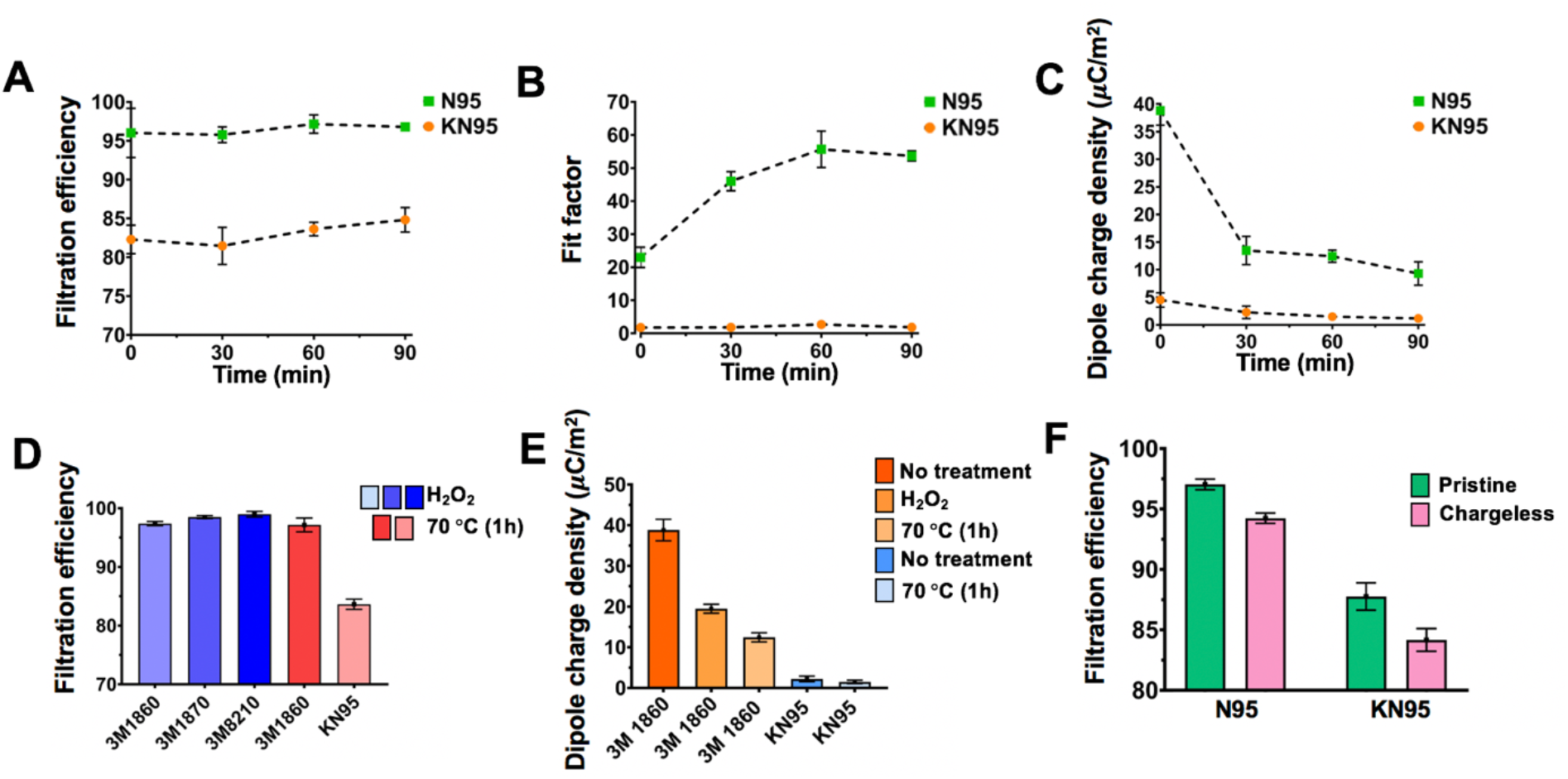
Comparison of N95 and KN95. Changes of (A) filtration efficiency (B) fit factor (C) dipole charge density of N95 and KN95 respirators after each cycle of 70 °C treatment. (D) Filtration efficiency of N95 respirators (3M 1860, 3M 1870, 3M 8210) disinfected by VHP method compared with 3M 1860 and KN95 disinfected by 70 °C heat treatment. (E) The decrease of dipole charge density is shown in the 3M 1860 disinfected by VHP method and 70 °C heat treatment. (F) Filtration efficiencies between the pristine and chargeless respirators. Dipole charges were removed by isopropanol (IPA) solution. Error bars represent the standard deviations of 5 measurements.

Changes of dipole charge density after each cycle of 70 °C treatments on N95 and KN95 are shown in **Figure 3C**. For the pristine materials, the N95 had 9-fold higher dipole charge density (38.82 μC/m^2^) at the baseline compared to the KN95. It dropped by 67 percent (12.48 μC/m^2^) after 30 minutes of 70 °C treatment. The dipole charge density of KN95 similarly decreased but the baseline was much lower (4.6 μC/m^2^ for pristine material) and it dropped by 50 percent (2.27 μC/m^2^) after 30 minutes of 70 °C treatment. It is because higher initial dipole charge density will induce higher inner electrical field in the material, which results in a faster decay rate of dipole charge density in N95 at the first cycle of heat treatment.^37-39^ However, the dipole charge densities of N95 respirator were still at least 3-fold higher than the KN95 (**Figure 3C**).

The subsequent decrease of dipole charge density after each cycle of heat treatment on both respirators can be explained by two reasons. First, the dipolar charges in the polymer material depend on steady state trapping/detrapping. The detrapping coefficient of electrons and holes is described as *D* = *v* * exp (−*w*/*kbT*)where *v* is the attempt to escape frequency and *w* is the detrapping barrier.^39^ With the increasing of temperature (*T*), trapped electrons and holes will be easier to be detrapped. Second, charge transport at the interface between solid polymer and the air at the boundary follows Schottky law, which indicates the charge dissipation fluxes become larger at higher temperature.^37-39^

The moisture in humid air acted as a natural conductor that allowed dipole charges to dissipate.^40^ Thus, the dipole charge density of N95 decreased to 19.48 μC/m^2^ after H_2_O_2_ treatment due to the involvement of moisture. The dipole charge was 12.48 μC/m^2^ after 60 minutes of 70 °C treatment (**Figure 3E**). Despite such differences in dipole charge density, there were no significant changes in filtration efficiency between H_2_O_2_ (97.38 %) and heat treatment (97.15 %) on the N95 respirator (**Figure 3D**) because of the involvement of mechanical filtration^34^ and the particle size.^36^ To minimize the effect of electrostatic attraction,^41^ we used isopropanol (IPA) to remove all the dipole charges in the filter layer and measured filtration efficiency which correspond to **Figure 3F**. The filtration efficiency of the N95 respirator decreased from 97.03 % to 94.24 % after removal of all the dipole charge; however, mechanical filtration and particle size were still leading to good filtration.

We next conducted 150 °C heat treatment as a positive control of the 70 °C, and fiber deformation occurred at the inner layer of N95 and inner/cotton layer of KN95 (**Figure 4**). The spun-bond PP of N95 started to melt at 150 °C, leading to fiber linkages with other fibers nearby (**Figure 4B**). Spun-bond PP of KN95 began to break their bonds (**Figure 4D**) showing their lower thermal durability compared to N95s. In addition, there were balloon-shaped fiber expansions at the cotton fibers of KN95 (**Figure 4F**). Cotton fibers consist of 95% cellulose and 0.6% natural wax, which serves as an outer layer.^42^ Those balloon-shaped fibers were caused by the expansion of the outer shell of cotton fiber and the inner core of cotton fiber kept maintaining its structure (**Figure S2**). Since the filter layer made of melt-blown PP were resistant to high temperature,^13^ there was no damage observed from the filter layer of both N95 and KN95 respirators (**Figure S3**). Those structural deformations occurred at high temperature and demonstrated that 70 °C was a suitable temperature for dry-heat method.

**Figure 4.**
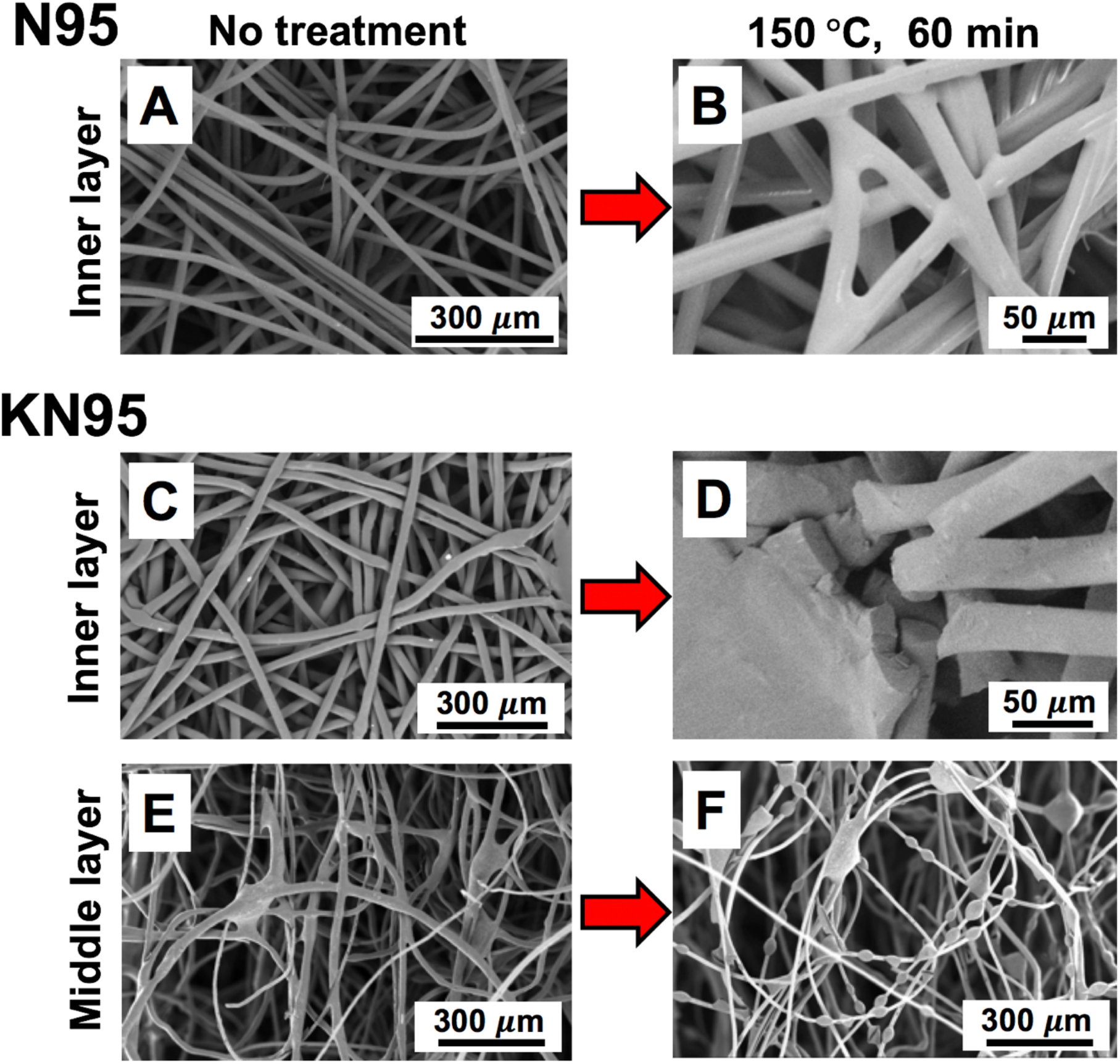
Fiber deformation at 150 °C treatment. (A) Inner layer of N95 (no treatment) (B) Melting happens at the fiber intersections. (C) Inner layer of KN95 (no treatment) (D) fiber fracture occurs after 150 °C treatment. (E) Middle layer of KN95 (no treatment) (F) Balloon-shaped fiber expansions happen after 150 °C treatment.

## Conclusion

In this study, we investigated whether KN95 is interchangeable with N95 respirators. We compared their baseline features as well as their physical filtration properties after disinfection via a dry-heat method chosen for its practicability, non-toxicity, and accessibility. Compared to the initial conditions, the filtration efficiencies of KN95 and N95 respirators increased after heat treatments; however, the filtration efficiencies stayed within a certain range after 60 minutes of heat treatment rather than a steady rise. The filtration efficiency as well as fit factor of N95 respirator were still much higher than KN95 even after heat treatments. In addition, structural instabilities such as fracture and balloon-shaped fiber expansion were found in KN95 respirators when heated at 150 °C.

Dipole charge density was also investigated in this study because electrostatic charge is involved in the filtration performance of respirators. Although the decreases in dipole charge density were observed in both respirators after several heat treatments, there was no quantitative relationship between dipole charge density and filtration efficiency since multiple parameters were involved in determining filtration efficiency. However, the dipole charge density of KN95 is at least 10-fold lower than the N95s. Finally, KN95 respirators are not replaceable with N95 respirators for healthcare workers due to their lower filtration efficiency, fit factor, and dipole charge density before and after heat treatment.

## Materials and Methods

### Sample preparation

N95 (3M 1860) and KN95 (Yomasi) respirators were used in this study. The N95 is NIOSH-approved with 95% filtration efficiency.^43^ The KN95 is GB 2626-2019-approved with 95% filtration efficiency.^44^ Each sample was cut into 2 cm ×2 cm before putting it into the oven. After each cycle of heat treatment, it was stored in the 50 mL conical centrifuge tube. N95 respirators (3M 1860, 3M 1870, 3M 8210) sterilized by VHP method were obtained from UCSD School of Medicine (Sandiego, CA, USA).

### Dry-heat treatment

A thermostat-controlled heating oven (ThermoFisher) was used for heat treatment. The interior size of the oven is 34. 3 cm (length), 35.4 cm (width), and 50.8 cm (height) for 62 L total volume. There were three shelves in the oven where respirators could be placed without stacking them together. Thus, it was capable of heating 18 respirators at once. The oven could manually control temperature range up to 330 ° C, and the fan in the oven maintains dry-heat condition. Contaminated N95 and KN95 respirator were put in the oven at 70 °C which can inactivate SARS-CoV-2.^23, 32^ We conducted 150 °C treatment as a positive control because operating temperature for polypropylene is 90 °C.^8^

### Fit factor & Material filtration test

We used a PortaCount Plus Model 8020 respirator fit tester which can measure the number of particles pre- and post-filtration per cubic centimeter (cc) along with providing a fit factor. The machine is best used in a closed room where the particle count is at least 30,000 particles per cc. All measurements were taken in the presence of two lit candles in a closed room to make sufficient particles in the air. The fit factor was taken by using a test probe kit which consists of a metal probe and a metal pin. The probe was placed on the metal pin and the pin was used to break through the KN95 or N95 respirator material in turn placing the probe within the respirator material and creating an airtight channel (**Figure S4A**). The machine’s air inlet tubing was attached to the probe after an individual wore the respirator (**Figure S4C**). The machine calculated fit factor by using the following equation:^45^

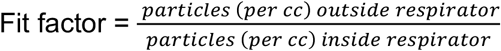

For material filtration tests, N95 and KN95 respirators were cut into 2 cm squares and placed into a 2 cm diameter cylindrical chamber with a swivel barb outflow adapter and an inflow tubing from the ambient atmosphere (Fig. S4B). When the material was placed in the chamber, the outflow swivel barb adapter was connected to the inflow tubing of the machine (Fig. S4D). The number of particles going to the PortaCount was measured for 30 second. Filtration efficiency of the sample is calculated using the following equation:^46^

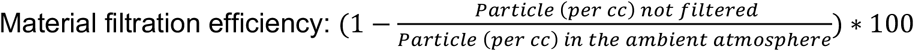

### Electron microscopy imaging

A scanning electron microscope (SEM; FEI Apreo) was used to examine the morphology of spun-bond and melt-blown PP fibers. SEM images were taken at an accelerating voltage of 1 kV and a current of 0.10 nA. The diameter of each fiber was calculated by ImageJ ^47^.

### Dipole charge density

A Trek model 344 electrostatic voltmeter was then used to measure the dipolar surface voltage of the filter layer. During the voltage measurement, one side of the filter layer was attached onto a grounded plate electrode, and a non-contact Kelvin probe was held 5 mm above the sample’s top surface along its center line to calculate surface charge. The dipole charge density was calculated based on Poisson equation.^48^

### Isopropanol (IPA) treatment

Filter layer of N95 and KN95 respirators was dipped into IPA solution for 30 second to remove all of the dipole charges. Then, the samples were dried at 35 °C in the air for 1 hour.

## Data Availability

The authors confirm that the data supporting the findings of this study are available within the article and/or its supplementary materials.

## Supporting Information

SEM images of N95 respirator after the filtration test, A balloon-shaped fiber expansion happened in KN95 respirator, Fiber stability of the filter layer, and Fit factor & Filtration test of N95 respirator. The supporting information is available free of charge on the ACS Publication websites

## Notes

The authors declare no competing financial interest.

## Acknowledgement

The authors acknowledge funding from the University of California Tobacco Related-Disease Research Program (TRDRP) under Emergency COVID-19 Research Seed Funding award number R00RG2515. The authors also acknowledge NIH under grants DP2 HL137187 and R21 AG065776 and NSF under grant #1845683. Fig. 1 and Table of Contents were created from www.Biorender.com. We acknowledge Bevis Respirator Consultants for use of the PortaCount system.

## Table of Contents (TOC)

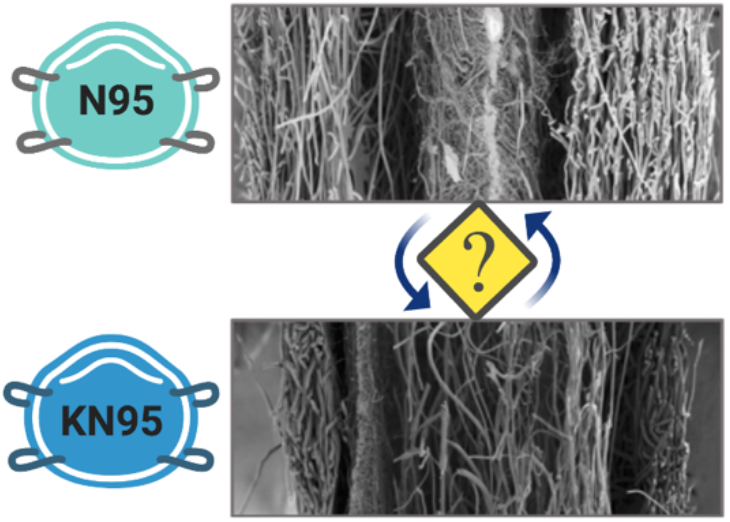

## References

(1) Baud, D.; Qi, X.; Nielsen-Saines, K.; Musso, D.; Pomar, L.; Favre, G. Real estimates of mortality following COVID-19 infection. The Lancet infectious diseases 2020.

(2) Yan, R.; Zhang, Y.; Li, Y.; Xia, L.; Guo, Y.; Zhou, Q. Structural basis for the recognition of SARS-CoV-2 by full-length human ACE2. Science 2020, 367 (6485), 1444–1448.

(3) Wang, J.; Du, G. COVID-19 may transmit through aerosol. Irish Journal of Medical Science (1971-) 2020, 1–2.

(4) Prather, K. A.; Wang, C. C.; Schooley, R. T. Reducing transmission of SARS-CoV-2. Science 2020.

(5) Qian, Y.; Willeke, K.; Grinshpun, S. A.; Donnelly, J.; Coffey, C. C. Performance of N95 respirators: filtration efficiency for airborne microbial and inert particles. American Industrial Hygiene Association Journal 1998, 59 (2), 128–132.

(6) Chawla, K. Fibrous materials, Cambridge university press: 2016.

(7) Konda, A.; Prakash, A.; Moss, G. A.; Schmoldt, M.; Grant, G. D.; Guha, S. Aerosol filtration efficiency of common fabrics used in respiratory cloth masks. ACS nano 2020, 14 (5), 6339–6347.

(8) Zerin, I.; Datta, E. A Review Article on Applications of Filter Cloth. Int. J. Cloth. Sci 2018, 5, 1–6.

(9) Gobi, N.; Evangelin, S.; Kasthuri, R. Multilayer nonwoven fabrics for filtration of micron and submicron particles. J Textile Eng Fashion Technol 2019, 5 (2), 81–84.

(10) Liu, J.; Zhang, X.; Zhang, H.; Zheng, L.; Huang, C.; Wu, H.; Wang, R.; Jin, X. Low resistance bicomponent spunbond materials for fresh air filtration with ultra-high dust holding capacity. RSC advances 2017, 7 (69), 43879–43887.

(11) Dutton, K. C. Overview and analysis of the meltblown process and parameters. Journal of Textile and Apparel, Technology and Management 2009, 6 (1).

(12) Mukhopadhyay, A. Composite nonwovens in filters: applications. In Composite Non-Woven Materials; Elsevier: 2014; pp 164–210.

(13) Brown, R. C. Air filtration: an integrated approach to the theory and applications of fibrous filters, Pergamon: 1993.

(14) Lee, Y.; Wadsworth, L. C. Structure and filtration properties of melt blown polypropylene webs. Polymer Engineering & Science 1990, 30 (22), 1413–1419.

(15) Pu, Y.; Zheng, J.; Chen, F.; Long, Y.; Wu, H.; Li, Q.; Yu, S.; Wang, X.; Ning, X. Preparation of polypropylene micro and nanofibers by electrostatic-assisted melt blown and their application. Polymers 2018, 10 (9), 959.

(16) Wang, C.-S. Electrostatic forces in fibrous filters—a review. Powder Technology 2001, 118 (1-2), 166–170.

(17) Matsuda, Y.; Saito, Y.; Tasaka, S. Dipole polarization formed on surface of polypropylene electrets. IEEE Transactions on Dielectrics and Electrical Insulation 2010, 17 (4), 1015–1020.

(18) Kravtsov, A.; Brünig, H.; Zhandarov, S.; Beyreuther, R. The electret effect in polypropylene fibers treated in a corona discharge. Advances in Polymer Technology: Journal of the Polymer Processing Institute 2000, 19 (4), 312–316.

(19) Ranney, M. L.; Griffeth, V.; Jha, A. K. Critical supply shortages—the need for ventilators and personal protective equipment during the Covid-19 pandemic. New England Journal of Medicine 2020, 382 (18), e41.

(20) Kim, T.; Zhang, Q.; Li, J.; Zhang, L.; Jokerst, J. V. A gold/silver hybrid nanoparticle for treatment and photoacoustic imaging of bacterial infection. ACS nano 2018, 12 (6), 5615–5625.

(21) Mantri, Y.; Davidi, B.; Lemaster, J. E.; Hariri, A.; Jokerst, J. V. Iodide-doped precious metal nanoparticles: measuring oxidative stress in vivo via photoacoustic imaging. Nanoscale 2020, 12 (19), 10511–10520.

(22) Ippolito, M.; Iozzo, P.; Gregoretti, C.; Cortegiani, A. Counterfeit filtering facepiece respirators are posing an additional risk to healthcare workers during COVID-19 Pandemic. American Journal of Infection Control 2020.

(23) Fischer, R. J.; Morris, D. H.; van Doremalen, N.; Sarchette, S.; Matson, M. J.; Bushmaker, T.; Yinda, C. K.; Seifert, S. N.; Gamble, A.; Williamson, B. N. Effectiveness of N95 Respirator Decontamination and Reuse against SARS-CoV-2 Virus. Emerging Infectious Diseases 2020, 26 (9).

(24) Sandle, T. Biocontamination Control for Pharmaceuticals and Healthcare, Academic Press: 2018.

(25) Food, U.; Administration, D., Final report for the Bioquell hydrogen peroxide vapor (HPV) decontamination for reuse of N95 respirators. White Oak, MD: FDA; 2016.

(26) Xiang, Y.; Song, Q.; Gu, W. Decontamination of Surgical Face Masks and N95 Respirators by Dry Heat Pasteurization for One Hour at 70° C. American Journal of Infection Control 2020.

(27) Liao, L.; Xiao, W.; Zhao, M.; Yu, X.; Wang, H.; Wang, Q.; Chu, S.; Cui, Y. Can N95 respirators be reused after disinfection? How many times? ACS nano 2020.

(28) Juang, P. S.; Tsai, P. N95 Respirator Cleaning and Reuse Methods Proposed by the Inventor of the N95 Mask Material. Journal of Emergency Medicine 2020.

(29) Cai, C.; Floyd, E. L. Effects of Sterilization With Hydrogen Peroxide and Chlorine Dioxide on the Filtration Efficiency of N95, KN95, and Surgical Face Masks. JAMA Network Open 2020, 3 (6), e2012099–e2012099.

(30) Ippolito, M.; Vitale, F.; Accurso, G.; Iozzo, P.; Gregoretti, C.; Giarratano, A.; Cortegiani, A. Medical masks and Respirators for the Protection of Healthcare Workers from SARS-CoV-2 and other viruses. Pulmonology 2020.

(31) Grafe, T.; Graham, K. Polymeric nanofibers and nanofiber webs: a new class of nonwovens. International Nonwovens Journal 2003, (1), 1558925003os–1200113.

(32) Chin, A.; Chu, J.; Perera, M.; Hui, K.; Yen, H.-L.; Chan, M.; Peiris, M.; Poon, L. Stability of SARS-CoV-2 in different environmental conditions. medRxiv 2020.

(33) Frederick, E. R. Fibers, electrostatics, and filtration: A review of new technology. Journal of the Air Pollution Control Association 1980, 30 (4), 426–431.

(34) Mostofi, R.; Wang, B.; Haghighat, F.; Bahloul, A.; Jaime, L. Performance of mechanical filters and respirators for capturing nanoparticles−Limitations and future direction. Industrial health 2010, 48 (3), 296–304.

(35) Kim, C. S.; Bao, L.; Okuyama, K.; Shimada, M.; Niinuma, H. Filtration efficiency of a fibrous filter for nanoparticles. Journal of nanoparticle research 2006, 8 (2), 215–221.

(36) He, X.; Reponen, T.; McKay, R. T.; Grinshpun, S. A. Effect of particle size on the performance of an N95 filtering facepiece respirator and a surgical mask at various breathing conditions. Aerosol Science and Technology 2013, 47 (11), 1180–1187.

(37) Alison, J.; Hill, R. A model for bipolar charge transport, trapping and recombination in degassed crosslinked polyethene. Journal of Physics D: Applied Physics 1994, 27 (6), 1291.

(38) Baudoin, F.; Le Roy, S.; Teyssedre, G.; Laurent, C. Bipolar charge transport model with trapping and recombination: an analysis of the current versus applied electric field characteristic in steady state conditions. Journal of Physics D: Applied Physics 2007, 41 (2), 025306.

(39) Le Roy, S.; Teyssedre, G.; Laurent, C.; Montanari, G. C.; Palmieri, F. Description of charge transport in polyethylene using a fluid model with a constant mobility: fitting model and experiments. Journal of Physics D: Applied Physics 2006, 39 (7), 1427.

(40) Chen, L.; Shi, Q.; Sun, Y.; Nguyen, T.; Lee, C.; Soh, S. Controlling surface charge generated by contact electrification: Strategies and applications. Advanced Materials 2018, 30 (47), 1802405.

(41) Ohmi, T.; Sudoh, S.; Mishima, H. Static charge removal with IPA solution. IEEE Transactions on semiconductor manufacturing 1994, 7 (4), 440–446.

(42) Wakelyn, P. J.; Bertoniere, N. R.; French, A. D.; Thibodeaux, D. P.; Triplett, B. A.; Rousselle, M.-A.; Goynes Jr, W. R.; Edwards, J. V.; Hunter, L.; McAlister, D. D. Cotton fiber chemistry and technology, CRC Press: 2006.

(43) Zhuang, Z.; Coffey, C. C.; Ann, R. B. The effect of subject characteristics and respirator features on respirator fit. Journal of Occupational and Environmental Hygiene 2005, 2 (12), 641–649.

(44) Rubio-Romero, J. C.; del Carmen Pardo-Ferreira, M.; García, J. A. T.; Calero-Castro, S. Disposable masks: Disinfection and sterilization for reuse, and non-certified manufacturing, in the face of shortages during the COVID-19 pandemic. Safety Science 2020, 104830.

(45) Janssen, L. L.; Luinenburg, M. D.; Mullins, H. E.; Nelson, T. J. Comparison of three commercially available fit-test methods. AIHA Journal 2002, 63 (6), 762–767.

(46) Rengasamy, S.; Shaffer, R.; Williams, B.; Smit, S. A comparison of facemask and respirator filtration test methods. Journal of occupational and environmental hygiene 2017, 14 (2), 92–103.

(47) Rasband, W. S., ImageJ. Bethesda, MD: 1997.

(48) Kou, R.; Zhong, Y.; Qiao, Y. Flow electrification of corona-charged polyethylene terephthalate film. arXiv preprint arXiv:2005.14385 2020.

